# Trends in Adolescent Pregnancy and Early ANC: Implications for Cervical Cancer Prevention in Zambia

**DOI:** 10.1101/2025.06.13.25329565

**Authors:** Newton Nyirenda, Whiteson Mbele, Chabota Simweemba

**Author notes:** **Corresponding Author** Newton Nyirenda, MD, MSc.

## Abstract

**Background:** Early initiation of antenatal care (ANC) is a cornerstone of maternal and child health, enabling timely detection of complications and delivery of preventive interventions, including education about cervical cancer. However, coverage of early ANC remains uneven across low-resource settings. This study assessed patterns and predictors of early antenatal care initiation in Zambia by analysing data from the 2007, 2013/14, and 2018 Demographic and Health Surveys, with an emphasis on equity-related disparities.

**Methods:** We analyzed pooled cross-sectional DHS data from 20,548 women aged 15–49 who had a recent live birth and received at least one ANC visit. Early ANC was defined as the first visit occurring during the first trimester. We generated weighted descriptive statistics, plotted trends over time, and used multivariable logistic regression to identify factors associated with early ANC initiation. Analyses accounted for complex survey design.

**Results:** Early ANC coverage rose from 19.7% in 2007 to 37.2% in 2018. Adolescents (15–19 years) made up 9.2% of the sample and had lower early ANC rates (26.3%) compared to older women (28.4%). Multivariable models revealed a consistent rise in early ANC over time, with each subsequent survey year associated with increased odds (AOR: 1.09; 95% CI: 1.08– 1.11; p < 0.001). Older maternal age (AOR: 1.16; 95% CI: 1.01–1.32; p = 0.039), higher educational attainment (AOR: 1.06; 95% CI: 1.00–1.13; p = 0.049), and greater household wealth (AOR: 1.05; 95% CI: 1.01–1.09; p = 0.039) were all associated with significantly greater odds of initiating ANC early. Urban residence was linked to decreased odds (AOR: 0.71; 95% CI: 0.63–0.80; p < 0.001).

**Conclusion:** While early ANC coverage has improved in Zambia, gaps persist across age, education, wealth, and residence. Targeted interventions addressing adolescent needs and integrating cervical cancer prevention into ANC services may enhance timely care and promote health equity.

## INTRODUCTION

Adolescent pregnancy remains a pressing public health challenge, particularly in low- and middle-income countries (LMICs), where the incidence continues to be disproportionately high. In sub-Saharan Africa, adolescent fertility rates remain among the highest worldwide, with 101 births per 1,000 girls aged 15–19 recorded in 2021, more than double the global average of 41 per 1,000 births. This pattern reflects deeply rooted socioeconomic and cultural disparities that continue to shape adolescent reproductive outcomes (1,2). In Zambia, the adolescent fertility rate has shown persistent elevation over the past decades, with a 2018 Demographic and Health Survey (DHS) reporting that nearly 29% of girls aged 15–19 had begun childbearing (3). Early pregnancies are associated with an array of adverse outcomes, including maternal mortality, neonatal complications, and curtailed educational and economic opportunities for young women (4–6).

Antenatal care (ANC) is a critical component of maternal and newborn health, providing opportunities for timely screening, detection of pregnancy-related complications, and linkage to broader health and social services (7). Among adolescents, timely initiation of ANC, ideally during the first trimester, has been associated with improved birth outcomes and enhanced maternal wellbeing (8). Despite this, adolescents are consistently found to access ANC later than recommended, or attend fewer than the four visits advised by WHO guidelines. Barriers to timely ANC among young people include stigma, lack of autonomy, limited knowledge, and structural challenges such as distance to facilities and inadequate youth-friendly services(9,10).

Early ANC initiation also serves as a strategic entry point for delivering broader preventive services, including cervical cancer screening and education on human papillomavirus (HPV) vaccination, particularly in settings where adolescent girls have minimal contact with formal health systems (11). The World Health Organization emphasizes the integration of cervical cancer prevention into existing maternal and adolescent health platforms as a high-impact strategy for reducing future cancer burden (12). This approach is especially relevant in Zambia, where cervical cancer remains the leading cause of cancer-related mortality among women and is largely preventable through timely screening and HPV vaccination (13).

While previous studies have explored determinants of adolescent pregnancy and barriers to ANC uptake independently, few have examined national-level trends in the early initiation of ANC among adolescent mothers and the implications for preventive health interventions, particularly in the context of cervical cancer (14–16). Moreover, longitudinal assessments using standardized, population-based data such as DHS surveys offer an opportunity to evaluate progress over time, identify persistent disparities, and inform health system adaptations to better serve vulnerable groups.

This study aims to examine trends in early ANC initiation among adolescent mothers in Zambia using DHS data from 2007, 2013–14, and 2018. We further investigate whether disparities in early ANC uptake persist across sociodemographic strata and explore the implications of our findings for cervical cancer prevention policies. In doing so, this paper contributes to the evidence base supporting integrated adolescent reproductive health services and underscores the importance of early engagement with the health system in mitigating downstream risks, including cervical cancer.

## METHODS

### Study Design

This study employed a repeated cross-sectional design using data from the Zambia Demographic and Health Surveys (ZDHS) conducted in 2007, 2013/14, and 2018. The DHS employs a multistage stratified sampling design to obtain nationally representative data on demographic and health indicators. We focused on women aged 15–49 years who had a live birth in the five years preceding each survey and who reported receiving at least one antenatal care (ANC) visit.

### Study Population

The analytic sample consisted of 20,548 eligible women, comprising 1,939 adolescents (aged 15–19 years) and 18,609 older women (aged 20–49 years). Sample sizes by survey year were 4,054 for 2007, 9,211 for 2013/14, and 7,283 for 2018.

### Outcome Variable

The primary outcome was early antenatal care (ANC) initiation, defined as attending the first ANC visit during the first trimester (≤12 weeks gestation), in accordance with WHO guidelines. A binary variable was created to capture this, coded as 1 for early initiation and 0 otherwise.

### Key Explanatory Variables

The main independent variables included survey year (2007, 2013/14, 2018), age group (adolescent [15–19 years] versus older women [20–49 years]), education level (categorized using DHS variable v106: no education, primary, secondary, or higher), wealth quintile (v190), and place of residence (v025, coded as urban versus rural).

### Data Management and Harmonization

To facilitate pooled analysis, variables across surveys were harmonized and a subset of women with recent births and at least one ANC visit was selected. Survey-specific ITN variables were accounted for in the code logic. DHS sample weights (v005), primary sampling units (v021), and strata (v022) were used in all analyses.

### Statistical Analysis

All analyses incorporated the complex survey design elements of the DHS, including stratification, clustering, and sampling weights, using the survey package in R. Descriptive statistics were used to estimate weighted proportions of early antenatal care (ANC) initiation overall, across survey years, and by key sociodemographic variables. To examine temporal and subgroup variations, we fitted a multivariable quasi-binomial logistic regression model with early ANC initiation as the outcome. Independent variables included survey year, maternal age group, educational attainment, household wealth quintile, and place of residence.

### Ethical Considerations

This study used publicly available, de-identified DHS datasets. Ethical approval for DHS surveys was obtained by the original data collectors from the relevant institutional review boards in Zambia and ICF International. Additional ethical clearance for this secondary analysis was not required.

## RESULTS

A total of 20,548 women aged 15–49 who had a recent birth and received at least one antenatal care (ANC) visit were included in the analysis across the 2007, 2013/14, and 2018 Zambia Demographic and Health Surveys (DHS). The largest proportion of respondents was from the 2013/14 survey (43.8%), followed by 2018 (32.2%) and 2007 (24.0%). The majority were adults aged 20 and above (90.8%), with adolescents (15–19 years) accounting for 9.2% of the sample as shown in Table 1.

**Table 1.** Sociodemographic Characteristics of Women Aged 15–49 Who Attended at Least One ANC Visit, Zambia DHS 2007–2018.

| Characteristic | Category | Sample size(n) | Proportion (%) |
| --- | --- | --- | --- |
| Survey Year | 2007 | 4,054 | 24.0% |
|  | 2013/14 | 9,211 | 43.8% |
|  | 2018 | 7,283 | 32.2% |
| Age Group | Adolescent | 1,939 | 9.2% |
|  | Older | 18,609 | 90.8% |
| Education Level | None | 2,114 | 10.2% |
|  | Primary | 10,989 | 53.5% |
|  | Secondary | 6,608 | 32.4% |
|  | Higher | 829 | 4.0% |
| Wealth Quintile | Lowest | 4,529 | 20.4% |
|  | Second | 4,277 | 20.4% |
|  | Middle | 4,100 | 20.4% |
|  | Fourth | 4,082 | 20.4% |
|  | Highest | 3,495 | 18.4% |
| Residence | Rural | 12,860 | 62.8% |
|  | Urban | 7,622 | 37.2% |

**Table 2.** Early ANC Coverage by Sociodemographic Characteristics Among Zambian Women with a Recent Birth, 2007–2018.

| Characteristic | Category | Early ANC Coverage (%) |
| --- | --- | --- |
| **Age Group** | Adolescent | 26.3 |
|  | Older | 28.4 |
| **Education Level** | None | 29.4 |
|  | Primary | 27.6 |
|  | Secondary | 27.2 |
|  | Higher | 41.9 |
| **Wealth Quintile** | Lowest | 30.5 |
|  | Second | 28.4 |
|  | Middle | 27.1 |
|  | Fourth | 24.3 |
|  | Highest | 30.9 |
| **Place of Residence** | Rural | 29.6 |
|  | Urban | 25.9 |

Educational attainment varied, with 53.5% of participants having completed primary education, 32.4% reaching secondary education, and 4.0% attaining higher education. About one in ten women (10.2%) reported no formal education. Wealth distribution was relatively uniform across quintiles, reflecting an even representation of economic backgrounds. In terms of residence, 62.8% of women lived in rural areas, while 37.2% were urban residents.

**Figure 1.**
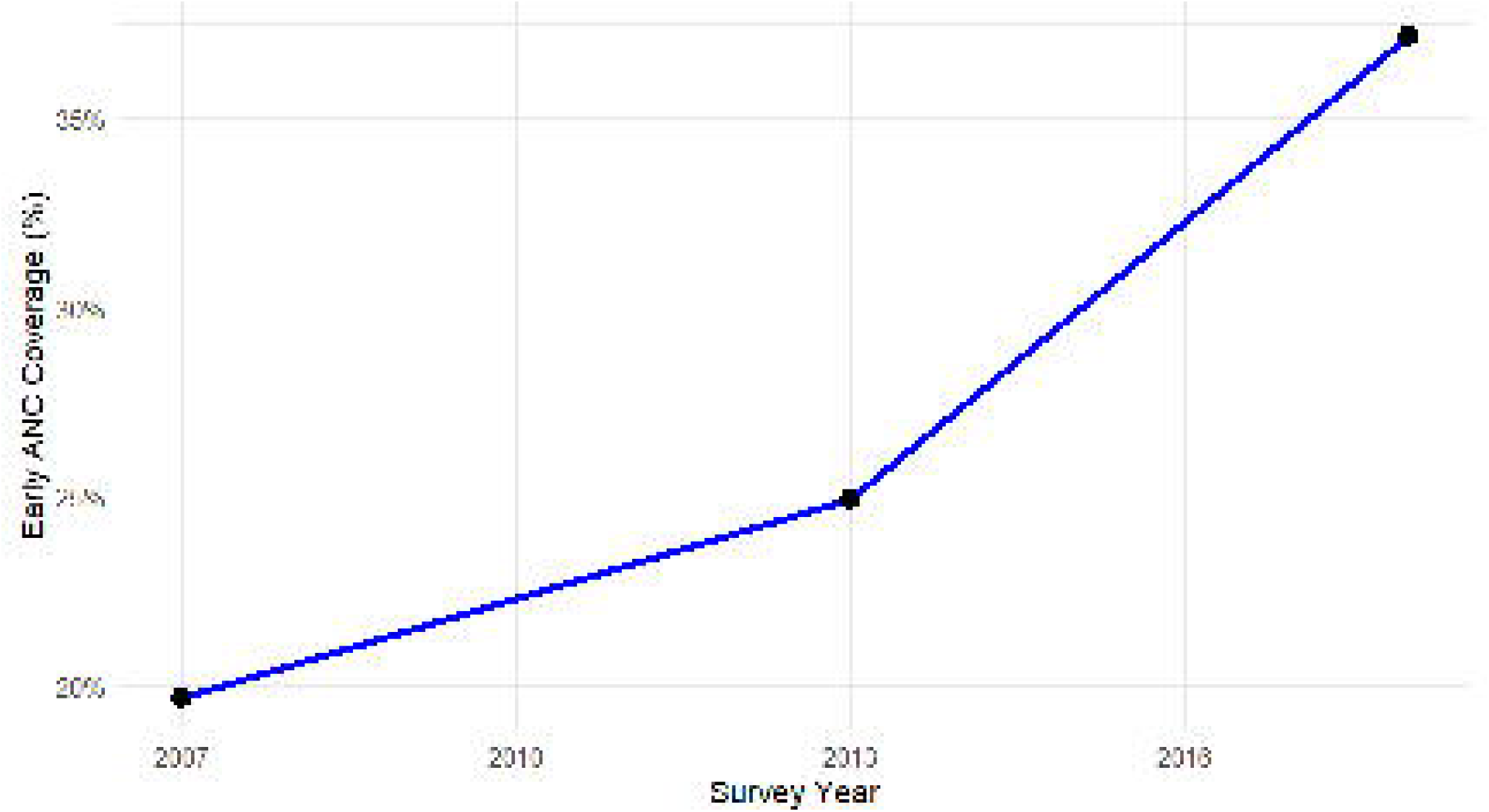
Trends in early antenatal care (ANC) coverage among Zambian women aged 15–49 years across three Demographic and Health Survey (DHS) rounds (2007, 2013/14, and 2018).

The proportion of women initiating ANC early increased steadily over the years, from 19.7% in 2007 to 37.2% in 2018.

Early initiation of antenatal care (ANC), defined as attending a first ANC visit within the first trimester, varied by sociodemographic characteristics. Coverage was slightly lower among adolescents (26.3%) compared to older women (28.4%). Women with higher education had the highest coverage (41.9%) relative to those with no education (29.4%) or primary education (27.6%). By wealth quintile, early ANC coverage ranged from 24.3% among those in the fourth quintile to 30.9% among those in the highest quintile. Urban residents had slightly lower early ANC coverage (25.9%) compared to rural residents (29.6%).

Multivariable logistic regression results presented in Table 3 show that survey year was significantly associated with higher odds of early ANC initiation (AOR: 1.09; 95% CI: 1.08, 1.11), indicating a steady improvement over time. Compared to adolescents, older women had slightly higher odds of initiating ANC early (AOR: 1.16; 95% CI: 1.01, 1.32). Higher educational attainment was also associated with increased likelihood of early ANC use (AOR: 1.06; 95% CI: 1.00, 1.13), as was increasing wealth (AOR: 1.05; 95% CI: 1.01, 1.09). However, women residing in urban areas had significantly lower odds of early ANC compared to rural residents (AOR: 0.71; 95% CI: 0.63, 0.80).

**Table 3.** Adjusted Odds Ratios (AOR), 95% Confidence Intervals, and p-values for Early Initiation of Antenatal Care Among Women in Zambia, 2007–2018.

| Predictor | AOR | 95% CI | p-value |
| --- | --- | --- | --- |
| Survey Year | 1.09 | (1.08, 1.11) | <0.001 |
| Age Group: Older vs Adolescent | 1.16 | (1.01, 1.32) | 0.039 |
| Education Level | 1.06 | (1.00, 1.13) | 0.049 |
| Wealth Quintile | 1.05 | (1.00, 1.09) | 0.039 |
| Urban vs Rural | 0.71 | (0.63, 0.80) | <0.001 |

## DISCUSSION

This study examined trends and sociodemographic determinants of early antenatal care (ANC) initiation among Zambian women using nationally representative data from the 2007, 2013/14, and 2018 Demographic and Health Surveys (DHS). The findings demonstrate a significant and encouraging increase in early ANC uptake over time, rising from 19.7% in 2007 to 37.2% in 2018. Despite this progress, overall early ANC coverage remains below national and international targets, including the WHO recommendation that all women initiate ANC within the first trimester to optimize maternal and fetal outcomes (7).

The observed upward trend is likely attributable to strengthened maternal health policies and increased public awareness campaigns during this period. Zambia introduced its Safe Motherhood Action Groups (SMAGs) and scaled up health communication efforts that emphasized early ANC engagement (17,18). The introduction of free maternal healthcare and community-based interventions may have also played a role in removing financial and logistical barriers (19). Similar trends in early ANC coverage have been documented in other sub-Saharan African countries, including Tanzania (20), suggesting a regional shift toward earlier maternal health service utilization.

Age emerged as a significant determinant of early ANC initiation, with adolescents being less likely than older women to seek care in the first trimester. This aligns with evidence from multiple low- and middle-income countries, where adolescent mothers often face stigma, lack autonomy, or lack knowledge about ANC services (21). Tailored interventions that address social and structural barriers unique to adolescents are warranted to improve timely ANC initiation in this vulnerable subgroup.

Higher educational attainment was associated with greater odds of early ANC, consistent with previous literature indicating that education enhances health literacy, awareness of pregnancy risks, and autonomy in healthcare decision-making (22,23). Studies from Ethiopia, Ghana, and Kenya have consistently reported similar associations (8,24). Education likely serves as both a direct and indirect enabler, influencing access to information, economic empowerment, and interaction with health systems.

Similarly, wealth status was positively associated with early ANC, although the gradient was modest. This finding reflects well-established evidence that economic resources enable women to overcome financial constraints such as transportation costs and informal service fees (25,26). Wealthier women may also live closer to health facilities or reside in better-served urban areas, although in this study, urban residence was paradoxically associated with lower odds of early ANC. This contradicts some regional patterns but aligns with findings from recent studies in South Africa, which attribute urban disadvantages to overcrowded facilities, longer waiting times, and perceptions of poor quality in public urban clinics (27,28).

The finding that urban women were less likely than rural women to initiate ANC early calls for deeper investigation. It is possible that rural women benefited more directly from targeted community outreach programs, such as SMAGs or mobile clinics, which may have improved ANC access and education in remote areas (29). Additionally, rural cultural norms surrounding pregnancy disclosure may differ, potentially encouraging earlier disclosure and care-seeking in some communities.

This study contributes to the growing body of evidence on ANC timing in sub-Saharan Africa by offering insights specific to Zambia using a decade-long pooled dataset. A key strength is the use of large, nationally representative samples and the application of survey weights and multivariable models to account for complex survey design and confounding factors.

However, the study has limitations. The use of cross-sectional data precludes causal inference. Timing of ANC was self-reported and may be subject to recall or social desirability bias. Moreover, potentially influential variables such as partner support, cultural beliefs, and facility-level factors were not available in the DHS data and could further explain variations in ANC timing.

In conclusion, although early ANC coverage in Zambia has improved over time, significant disparities remain by age, education, wealth, and residence. Interventions to promote early ANC should prioritize adolescents, expand health education efforts, and address urban health system bottlenecks. Ensuring equitable and timely access to maternal health services is critical for reducing preventable maternal and neonatal morbidity and mortality.

## CONCLUSION

This study provides evidence of significant progress in the early initiation of antenatal care (ANC) among Zambian women between 2007 and 2018, with early ANC coverage nearly doubling over the period. Despite this improvement, early ANC uptake remains suboptimal, particularly among adolescents, urban residents, and women with lower educational attainment or in the poorest wealth quintiles. Our findings show persistent sociodemographic disparities in maternal healthcare access and reinforce the urgency of implementing equity-focused interventions.

Given the critical role of timely ANC in improving maternal and neonatal outcomes, and its relevance in supporting cervical cancer prevention through early screening and education, greater efforts are needed to ensure inclusive access. Special attention should be directed toward improving coverage among underserved subgroups, including adolescent girls and women in urban informal settlements. Interventions that simultaneously address both demand-side and supply-side barriers will be essential in accelerating progress toward universal maternal health coverage and reducing future cancer risk through integrated prevention strategies.

## Data Availability

The DHS datasets analyzed during the current study are publicly available from The DHS Program repository at: https://dhsprogram.com/data

## Declarations

### Ethics approval and consent to participate

This study is based on publicly available, de-identified data from the Zambia Demographic and Health Surveys (DHS) conducted in 2007, 2013/14, and 2018. Ethical approval for DHS surveys was obtained by ICF International and the Zambian Ministry of Health through their respective institutional review boards. This secondary analysis did not require additional ethical approval.

## Consent for publication

Not applicable. The manuscript does not contain any individual-level data, images, or videos requiring consent.

## Availability of data and materials

The DHS datasets analyzed during the current study are publicly available from The DHS Program repository at: https://dhsprogram.com/data/available-datasets.cfm. The authors used Zambia DHS datasets from 2007, 2013/14, and 2018 after obtaining data access approval.

## Authors’ contributions

NN and WM conceptualized the study, conducted data analysis, interpreted the findings, and wrote the manuscript. CS reviewed the analysis outputs, contributed to interpretation of findings, and critically revised the manuscript. All authors read and approved the final version of the manuscript.

## Acknowledgements

The authors gratefully acknowledge the Demographic and Health Surveys (DHS) Program and the Zambian Central Statistical Office for providing access to the datasets used in this study.

